# Knowledge, Awareness, and Prescribing Practices Regarding Sugar-Free Paediatric Liquid Medicines Among Healthcare Professionals in Uttarakhand: A Cross-Sectional Study

**DOI:** 10.64898/2026.04.15.26350902

**Authors:** Kumari Neha Jha, Kalpna Chaudhry, Nitin Khanduri

## Abstract

**Background:** Paediatric liquid medicines (PLMs) routinely contain sucrose to improve palatability, yet their cariogenic potential is well established. Healthcare professionals’ awareness and prescribing practices regarding sugar-free PLMs have received limited study in India, particularly in Uttarakhand.

**Methods:** A descriptive cross-sectional study was conducted among 500 healthcare professionals aged ≥25 years, using a pilot-tested structured questionnaire (Cronbach’s α = 0.85), administered online and in person across Uttarakhand districts (January–March 2024). After excluding 69 incomplete responses, 431 participants were analysed (response rate: 86.2%), comprising general medicine practitioners (49%, n = 211), paediatricians (27%, n = 116), and dental practitioners (24%, n = 104). Descriptive statistics and chi-square tests were applied (p < 0.05).

**Results:** Prescription decisions were primarily driven by child’s age and weight (58%), cost (40%), and pharmaceutical brand (37%). While 88% recognised PLM sweetness and 67% were aware of pH–dental harm links, only 20% associated PLMs with dental caries. Overall awareness of hidden sugars was 73%. Eighty-three percent knew of sugar-free alternatives (50% local availability), yet 80% found them less palatable and 85% costlier. Only 48% routinely provided oral health advice. A statistically significant association was found between specialty and sugar-free PLM awareness (p = 0.03), with dental practitioners recording the highest awareness (90%).

**Conclusions:** Healthcare professionals demonstrated variable levels of knowledge, attitudes, and practices regarding PLMs, with critical gaps in caries recognition (20%) and oral health counselling (48%). Despite high sugar-free PLM awareness, uptake is constrained by perceived cost and palatability barriers. Targeted continuing medical education and policy measures, including sucrose-free labelling promotion, are needed to improve paediatric oral health outcomes in Uttarakhand.

**KEY MESSAGES:**

1. Only 20% of healthcare professionals in Uttarakhand associated pediatric liquid medicines (PLMs) with dental caries, representing a critical knowledge gap despite 88% recognising their sweetness.
2. Overall awareness of hidden sugars in PLMs was 73%, yet only 48% routinely provided post-prescription oral health counsellingsubstantially below international benchmarks.
3. Eighty-three percent were aware of sugar-free PLM alternatives, but adoption was constrained by perceived inferior palatability (80%) and higher cost (∼10% premium, cited by 85%).
4. Dental practitioners demonstrated significantly higher sugar-free PLM awareness than general practitioners and pediatricians (p = 0.03), supporting the case for interprofessional oral health education in medical training.
5. Targeted continuing medical education (CME) and policy measuresincluding sucrose-free labelling mandates and institutional formulary inclusionare needed to convert awareness into prescribing practice change.

## INTRODUCTION

Healthcare professionals particularly paediatricians and general physicians significantly influence children’s oral health through their medication prescribing patterns.Healthcare professionals particularly paediatricians and general physicians significantly influence children’s oral health through their medication prescribing patterns.^1^ Paediatric liquid medicines (PLMs) are indispensable in managing childhood illnesses, yet the routine addition of sucrose to improve palatability poses well-documented risks to developing dentition.^2^ The cariogenic potential of sucrose-containing PLMs was first identified in 1953^3^ and subsequently corroborated by studies demonstrating significantly higher caries rates in children receiving long-term sweetened medications.^4,5^

Sucrose in PLMs creates a cariogenic oral environment through increased viscosity, prolonged salivary clearance, and fermentable substrate provision for acidogenic bacteria.^6^ Beyond caries, the acidic pH of many analgesic and antipyretic syrups can cause enamel erosion,^7^ and iron-containing or antibiotic preparations are associated with tooth staining.^8^ Children requiring prolonged PLM courses are at particularly elevated risk,^9^ making prescriber awareness a critical determinant of paediatric oral health outcomes.

Despite the availability of sugar-free alternatives formulated with xylitol, sorbitol, or saccharin, their adoption among prescribers remains limited by perceived cost premiums, inferior palatability, and restricted local availability.^10^ Studies from Saudi Arabia,^11^ Nigeria,^12^ Brazil,^13^ and India^14^ document variable practitioner knowledge and practice, but data from Uttarakhand characterised by heterogeneous geography, socioeconomic diversity, and variable healthcare access are absent from the literature. This study, conducted with major representation from AIIMS Rishikesh, Seema Dental College & Hospital (SDCH), and Uttaranchal Dental & Medical Research Institute (UDMRI), aims to comprehensively evaluate knowledge, attitudes, and prescribing practices regarding sugar-free PLMs and identify gaps amenable to targeted intervention.

## METHODS

### Study design and setting

A descriptive cross-sectional study was conducted across multiple districts of Uttarakhand from January to March 2024, following STROBE reporting guidelines.^15^ Healthcare professionals from tertiary institutions (AIIMS Rishikesh, SDCH, UDMRI) and semi-urban clinics and district hospitals in Dehradun, Haridwar, Nainital, and other districts were included.

### Participants

A total of 500 licensed healthcare professionals aged ≥25 years with at least one year of active clinical practice involving paediatric patients were approached through online (Google Forms) and in-person questionnaire administration. Practitioners in exclusively administrative roles were excluded. Of 500 approached, 69 responses were excluded for incompleteness or insufficient data, yielding a final analytical sample of 431 (response rate: 86.2%). The sample comprised general medicine practitioners (49%, n = 211), paediatricians (27%, n = 116), and dental practitioners (24%, n = 104). Institutional distribution: AIIMS Rishikesh (56%, n = 241), SDCH (21%, n = 90), UDMRI (18%, n = 78), and other facilities (5%, n = 22). All participants were aged 25 years and above; 52.7% (n = 227) were female and 47.3% (n = 204) were male. Geographic representation spanned Dehradun (56%), Haridwar (15%), Tehri Garhwal(8%) Nainital (7%), Almora (5%), Pithoragarh (4%), Champawat (3%), and other districts.

### Ethical considerations

Ethical approval for this study was waived by the Institutional Ethics Committee of Seema Dental College & Hospital (SDCH), Rishikesh, as it involved a minimal-risk, questionnaire-based survey conducted via online (Google Forms) and in-person methods among healthcare professionals, with no patient intervention. Informed consent was obtained from all participants prior to data collection. Participation was voluntary, and responses were kept confidential, with data handled securely and used solely for research purposes, in accordance with the Declaration of Helsinki.^16^

### Data collection instrument

The structured 20-item questionnaire covered five domains: demographics and practice profile; prescription preferences and patterns; knowledge of PLM properties and dental effects; awareness of sugar-free alternatives; and attitudes and counselling practices. It was adapted from Bradley and Kinirons^17^ and Bawazir et al.,^11^ with expert validation by four specialists in paediatric dentistry, clinical pharmacology, and public health. A pilot study of 20 practitioners confirmed reliability (Cronbach’s α = 0.85) and face validity.

### Statistical analysis

Data were analysed using SPSS version 27.0 (IBM Corp., Armonk, NY, USA). Descriptive statistics (frequencies and percentages) summarised demographic and response variables. Chi-square tests assessed associations between categorical variables, including specialty and institutional affliation versus knowledge and awareness outcomes, and practice location (categorised as urban, semi-urban, and rural districts) versus prescription form preference. Subgroup analyses were planned a priori to examine variations by speciality, institutional affiliation, and practice location. Statistical significance was set at p < 0.05.

## RESULTS

### Participant demographics

The final sample (n = 431) comprised general medicine practitioners (49%, n = 211), paediatricians (27%, n = 116), and dental practitioners (24%, n = 104). All participants were aged 25 years and above, with a slight female majority (52.7%, n = 227). By institutional affiliation: AIIMS Rishikesh (56%, n = 241), SDCH (21%, n = 90), UDMRI (18%, n = 78), and others (5%, n = 22). Full demographic and KAP data are presented in Table 1.

**Table 1:** Knowledge, Awareness, and Prescribing Practices Regarding Sugar-Free Paediatric Liquid Medicines Among Healthcare Professionals in Uttarakhand (n = 431)

| Variable | Category/<br>Response | n (%) | Interpretation |
| --- | --- | --- | --- |
| **Demographics** |  |  |  |
| **Specialty** | General medicine practitioners | 211 (49%) | Largest group |
|  | Pediatricians | 116 (27%) |  |
|  | Dental practitioners | 104 (24%) |  |
| **Age** | 25 years and above (all active practitioners) | 431 (100%) | Inclusion criterion |
| **Gender** | Female | 227 (52.7%) |  |
|  | Male | 204 (47.3%) |  |
| **Institution** | AIIMS Rishikesh | 241 (56%) | Major tertiary referral centre |
|  | Seema Dental College & Hospital (SDCH) | 90 (21%) |  |
|  | Uttaranchal Dental & Medical Research Institute (UDMRI) | 78 (18%) |  |
|  | Other facilities | 22 (5%) |  |
| **Knowledge** |  |  |  |
| **PLMs recognised as sweet** | Yes | 379 (88%) | High palatability awareness |
|  | No | 52 (12%) |  |
| **PLMs associated with dental caries** | Yes | 86 (20%) | Critical knowledge gap |
|  | No/unsure | 345 (80%) |  |
| **pH-dental harm awareness** | Yes | 289 (67%) | Moderate awareness |
| **Hidden sugars awareness (overall)** | Yes | 315 (73%) | Sucrose, fructose, glucose most cited |
| **PLMs cause tooth staining** | Yes | 203 (47%) | Iron/antibiotic preparations suspected |
| **Awareness and Attitudes** |  |  |  |
| **Aware of sugar-free PLMs** | Yes | 358 (83%) | Exceeds most global comparators |
|  | No | 73 (17%) |  |
| **Sugar-free PLMs locally available** | Yes | 216 (50%) | Below international benchmarks |
| **SF PLMs perceived as less palatable** | Yes | 345 (80%) | Key barrier to adoption |
| **SF PLMs perceived as costlier (~10%)** | Yes | 367 (85%) | Structural cost barrier |
| **Support universal SF PLM availability** | Yes | 272 (62%) | Strong regional endorsement |
| **Prescribing Practice** |  |  |  |
| **Oral health advice given post-PLM** | Yes | 207 (48%) | Gap vs 65% global benchmark |
|  | No | 224 (52%) |  |
| **Frequency of SF PLM prescribing** | Always | 129 (30%) |  |
|  | Often / Seldom / Never | 173 / 108 / 21 | 40% / 25% / 5% |
| **Parental requests for SF PLMs** | Yes | 151 (35%) | Growing consumer awareness |
|  | No | 280 (65%) |  |
| **Referral to paediatric dentist** | Always/<br>Sometimes | 256 (59%) |  |
|  | Seldom/Never | 175 (41%) |  |
| **Interest in CME (PLMs and oral health)** | Yes | 391 (90%) | Very high motivation |
|  | No | 40 (10%) |  |

*SF = Sugar-Free; PLMs = Paediatric Liquid Medicines; AIIMS = All India Institute of Medical Sciences; SDCH = Seema Dental College & Hospital; UDMRI = Uttarakhand Dental & Medical Research Institute; CME = Continuing Medical Education. Domain heading rows are shown in blue.*

### Prescription preferences

Prescription decisions were primarily driven by child’s age and weight (58%), cost (40%), and pharmaceutical brand (37%). Oral liquid formulations were preferred by 95% of respondents, with syrups dominant for children aged 1–6 years (96%). Tablets were used by 22.8%, typically for older children or specific indications. No tetracycline prescriptions for the paediatric cohort were reported.^18^

### Knowledge of PLM properties

Eighty-eight percent of participants recognised PLMs as sweet; however, only 20% associated them with dental caries. Sixty-seven percent were aware of pH-related enamel harm, and 47% associated PLMs with tooth staining.^8^ Dental practitioners demonstrated the highest caries awareness (30%) compared to paediatricians (20%) and general practitioners (15%), confirmed by chi-square analysis (p = 0.03).^19^

### Hidden sugar awareness and oral health counselling

Overall, 73% of participants were aware of hidden sugars in PLMs, with sucrose (35%), fructose (25%), and glucose (22%) most frequently identified.^20^ Despite this, only 48% routinely provided oral health advice following PLM prescriptionbelow the 65% reported by McVeigh and Kinirons^21^ and the 70% by Al-Jobair et al.,^22^ indicating a persistent practice–knowledge gap.

### Attitudes toward sugar-free PLMs

Eighty-three percent of participants were aware of sugar-free PLM formulations, with 50% reporting local availability. However, 80% perceived sugar-free formulations as less palatable and 85% noted a cost premium of approximately 10%.^23^ Despite these perceived barriers, 62% supported universal availability of sugar-free PLMs, and 70% reported prescribing them always (30%) or often (40%). Parental requests for sugar-free options were cited by 35% of respondents.^24^ Ninety percent expressed interest in CME on PLM-related oral health topics.

### Statistical associations

Chi-square analysis confirmed statistically significant associations between specialty and sugar-free PLM awareness (p = 0.03), with dental practitioners recording the highest awareness (90%);^25^ between practice location (urban/semi-urban/rural districts, treated as a categorical variable) and prescription form preference (p = 0.04);^26^ and between specialty and frequency of referral to a paediatric dentist (p = 0.04). No significant associations were found for gender or years of experience (p > 0.05).

## DISCUSSION

This study demonstrates that healthcare professionals in Uttarakhand exhibited variable levels of knowledge, attitudes, and practices regarding sugar-free PLMs. The 83% sugar-free PLM awareness exceeds rates reported in Saudi Arabia (65%)^11^ and Nigeria,^12^ but the 20% caries awareness is substantially lower than the 35% in Brazil,^13^ 40% in Nigeria,^12^ and 45% in South India,^27^ underscoring a regional knowledge deficit with meaningful implications for preventable paediatric dental disease.

The primacy of child’s age and weight and cost as prescribing drivers is consistent with practice in resource-limited institutional settings^28^ and reflects the socioeconomic profile of the cohort. The 79% syrup preference corroborates global trends driven by palatability needs and sucrose’s pharmaceutical multi-functionality as a preservative and thickener.^6^ The significant association between practice location and prescription form preference (chi-square, p = 0.04) highlights geographic heterogeneity in prescribing behaviour that targeted policy interventions must account for.^29^

The 48% oral health counselling rate is below international benchmarks^21,22^ and illustrates that knowledge does not automatically translate into clinical practice consistent with Michie’s behaviour change wheel framework, which identifies knowledge as necessary but insufficient for behaviour change.^30^ The 85% cost perception and 80% palatability perception represent structural barriers that require pharmaceutical policy responses: mandatory sucrose-free labelling demonstrated to be effective in the United Kingdom^10^ and Australia^25^ institutional formulary adoption, and government subsidy mechanisms have all shown positive effects in comparable contexts.^24^

The specialty effect (dental practitioners 90% vs. general practitioners 72% sugar-free awareness, p = 0.03) confirms that dental training provides a meaningful knowledge advantage and supports the integration of oral health modules into medical curricula.^26^ With 90% of respondents expressing interest in CME, institutions such as SDCH and UDMRI are well placed to lead regional interdisciplinary training programmes. The heavy representation of AIIMS Rishikesh (56%) and the convenience sampling approach limit generalisability to the broader Uttarakhand practitioner population; future studies should employ stratified random sampling across all districts and incorporate objective clinical audit data to validate self-reported practices.^13,15^

## CONCLUSIONS

Healthcare professionals in Uttarakhand demonstrated variable levels of knowledge, attitudes, and practices toward sugar-free PLMs. Critical gaps in caries recognition (20%) and oral health counselling (48%) are most pronounced among non-dental practitioners. While sugar-free PLM awareness (83%) is encouraging and exceeds several international benchmarks, structural barriers of cost and palatability constrain uptake. Interprofessional CME, sucrose-free labelling mandates, and formulary-level policies are required to bridge the awareness–practice gap and reduce the burden of PLM-associated dental caries in Uttarakhand’s paediatric population.

**Fig. 1.**
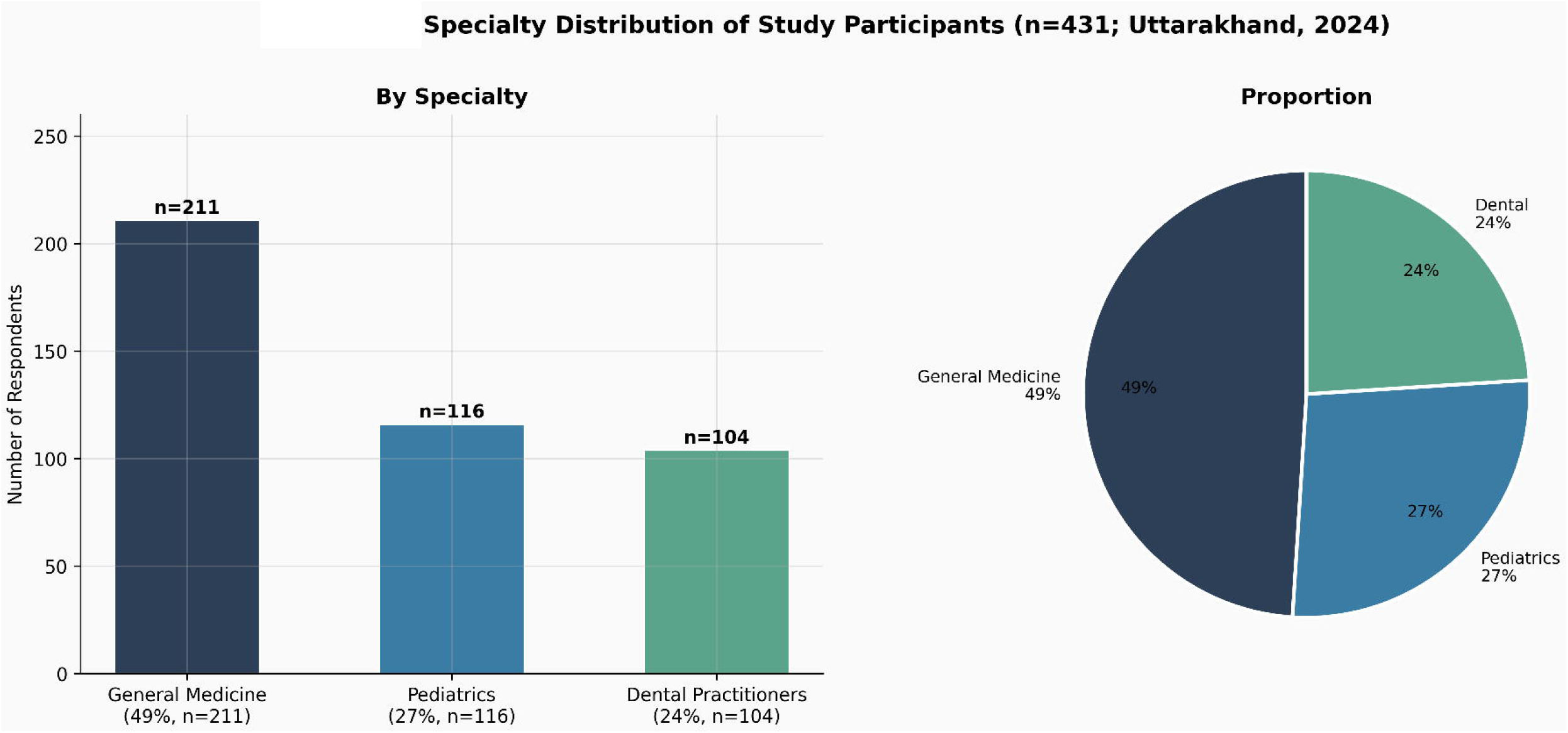
Specialty distribution of study participants (n = 431): bar chart and pie chart showing general medicine practitioners (49%, n = 211), paediatricians (27%, n = 116), and dental practitioners (24%, n = 104). Uttarakhand, India, 2024.

**Fig. 2.**
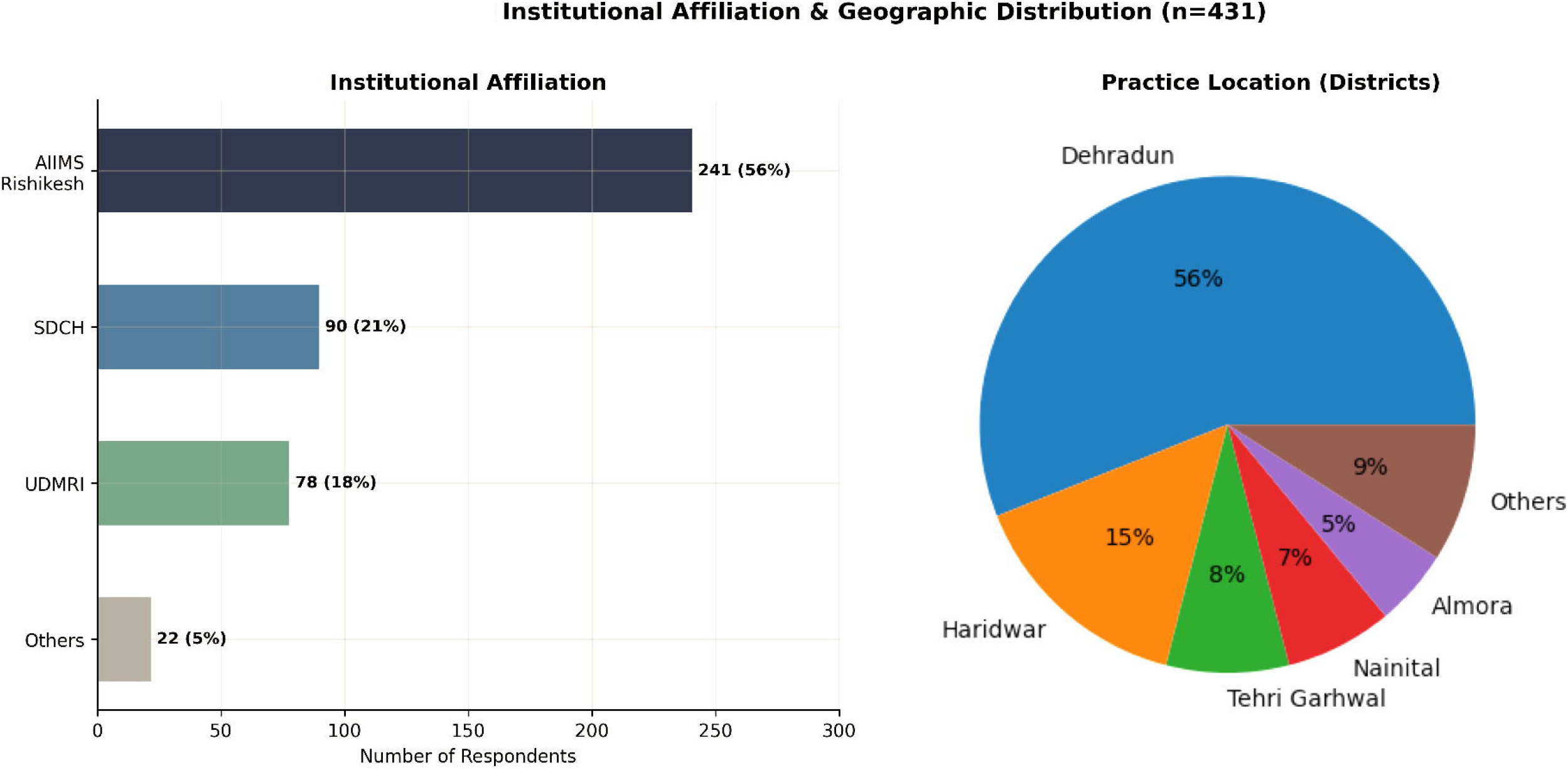
Institutional affiliation and district-level geographic distribution of respondents (n = 431): AIIMS Rishikesh 56% (n = 241), SDCH 21% (n = 90), UDMRI 18% (n = 78), others 5% (left panel); district-wise distribution pie chart (right panel).

**Fig. 3.**
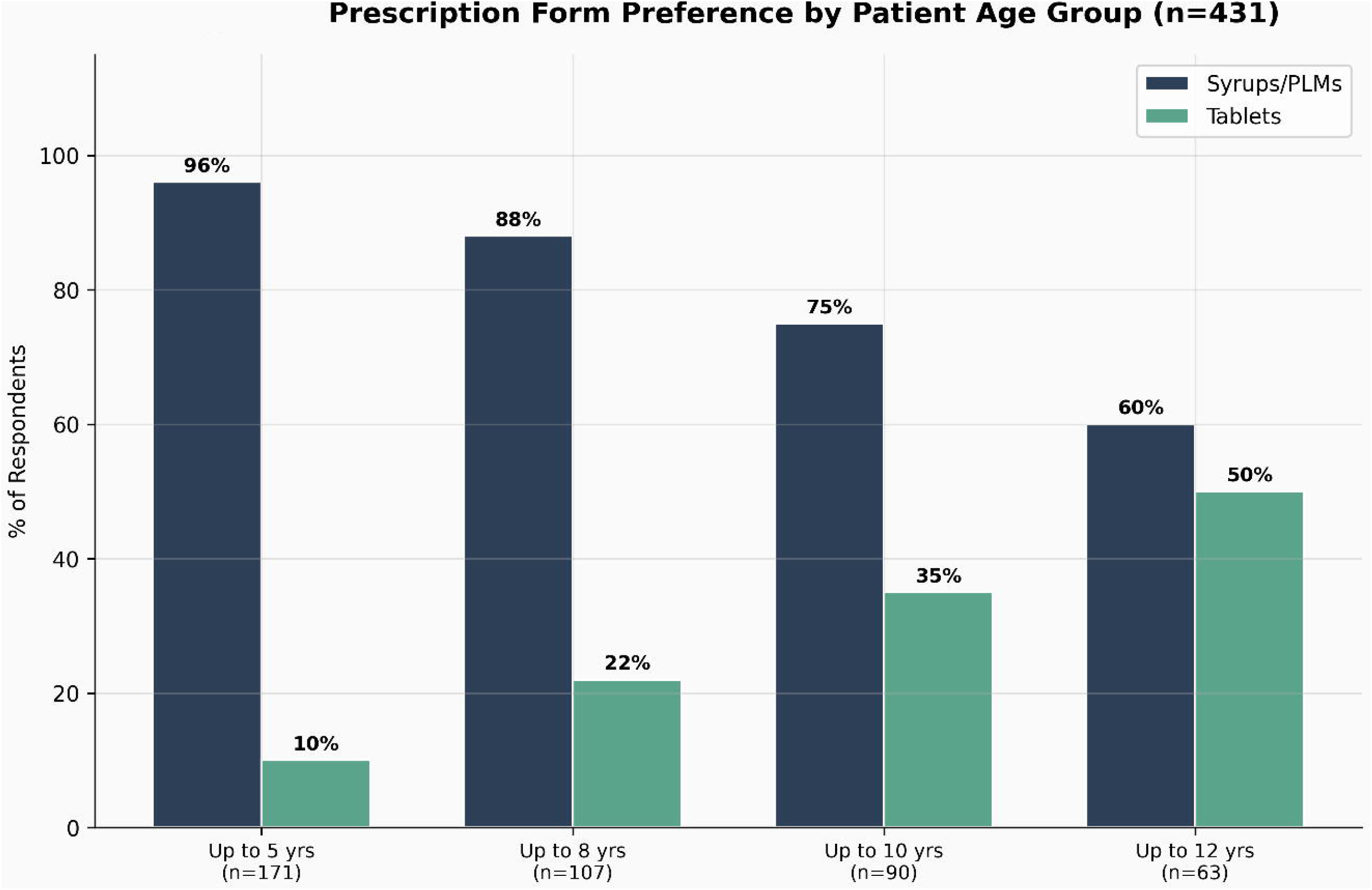
Prescription form preference by patient age group: grouped bar chart showing syrup/PLM and tablet prescribing rates for age groups up to 5, 8, 10, and 12 years (n = 431).

**Fig. 4.**
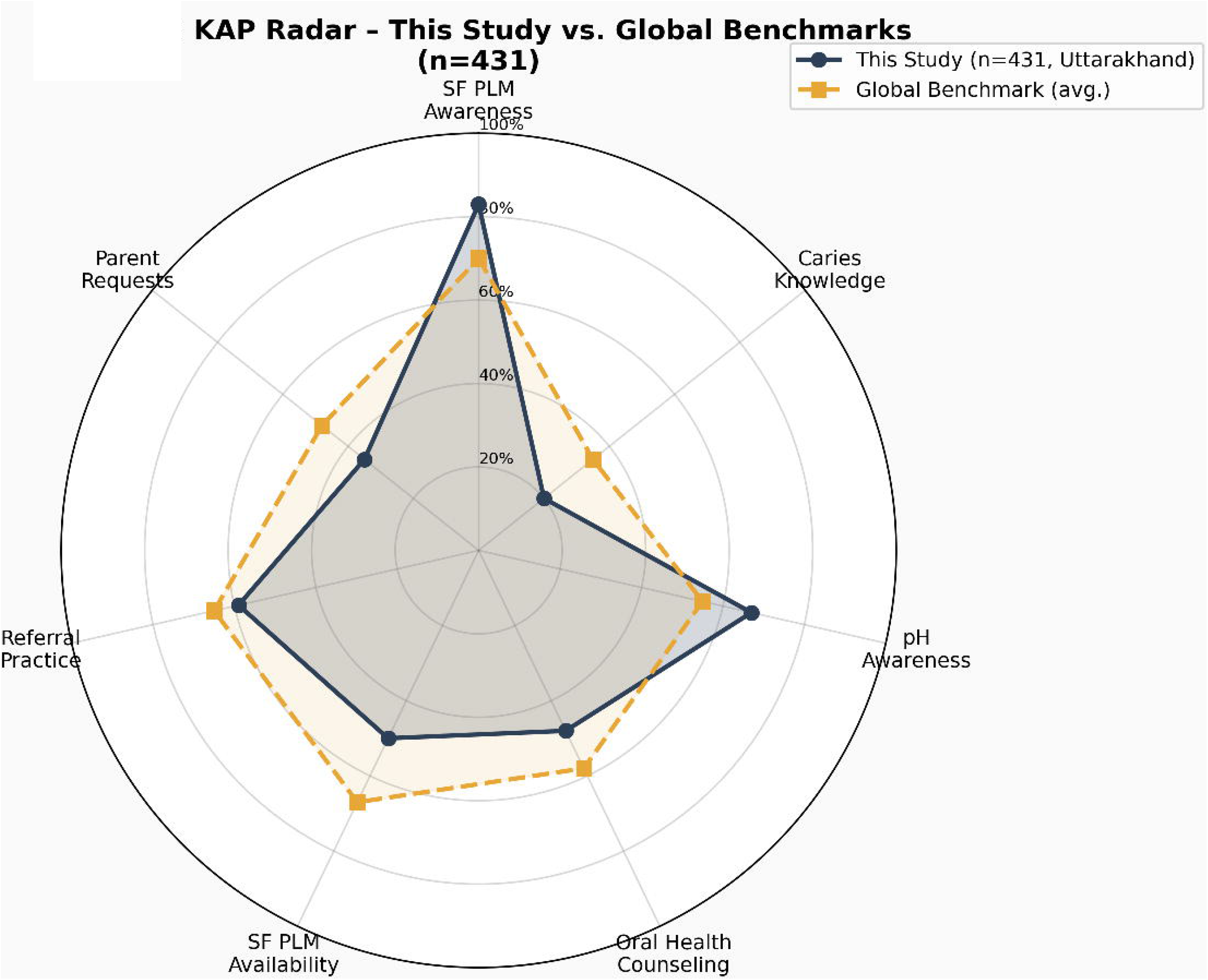
KAP radar profile comparing results of this study against global benchmark averages across seven outcome domains (n = 431).

**Fig. 5.**
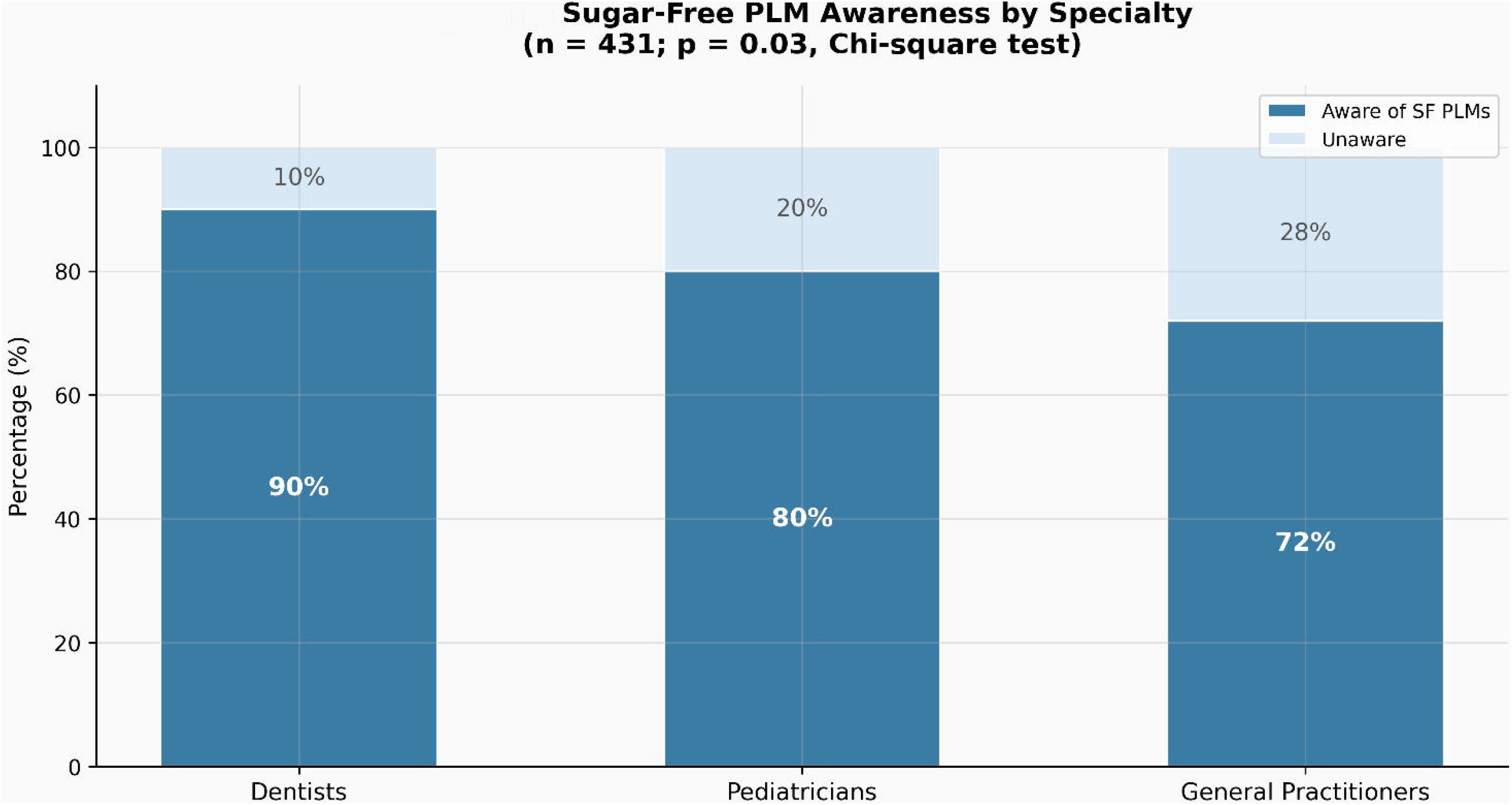
Sugar-free PLM awareness by specialty: stacked bar chart showing percentage aware and unaware among dental practitioners, paediatricians, and general practitioners (chi-square p = 0.03; n = 431).

**Fig. 6.**
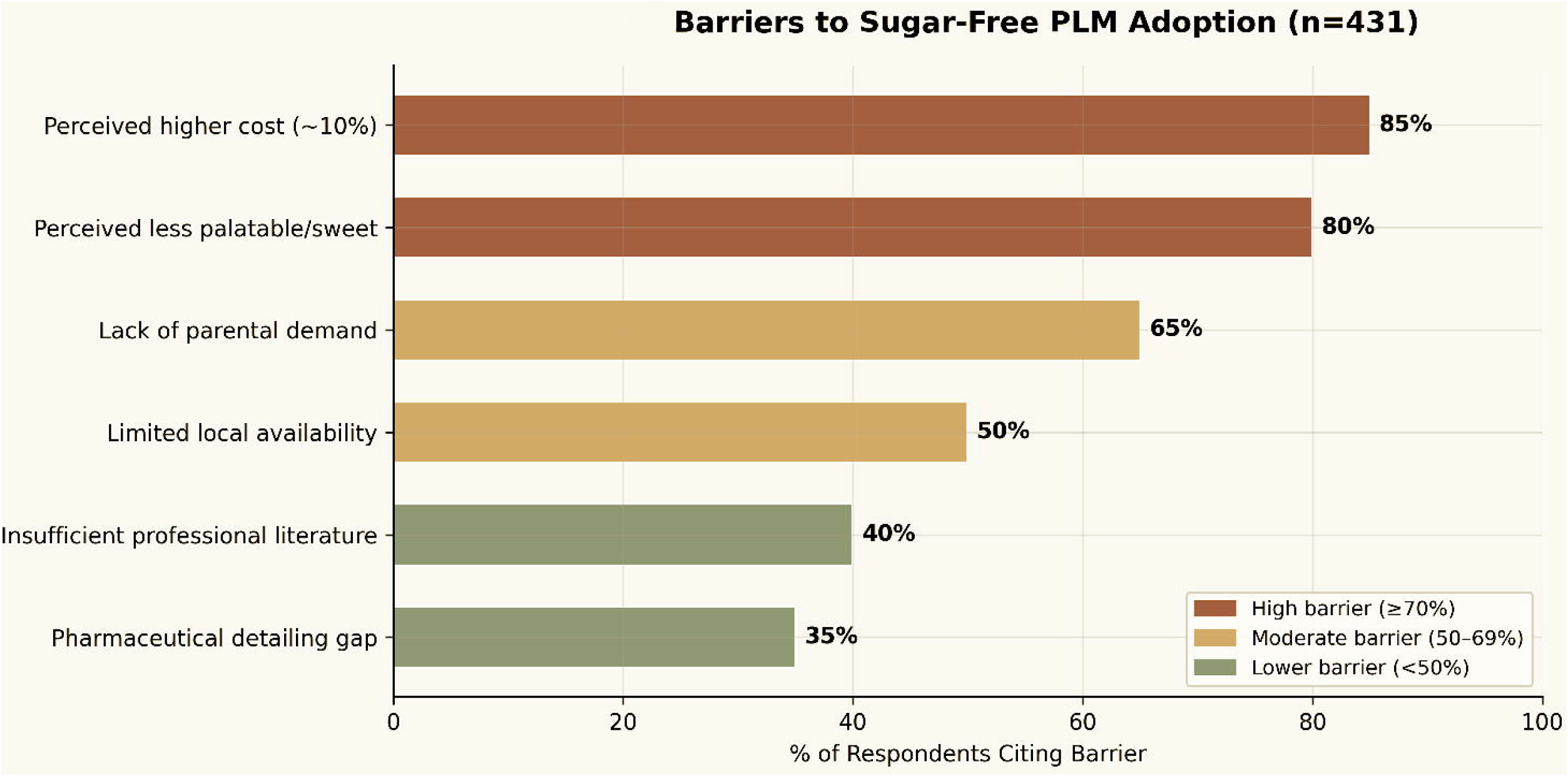
Key barriers to sugar-free PLM adoption as reported by respondents (n = 431): horizontal bar chart categorised by barrier severity.

## Data Availability

All data produced in the present study are available upon reasonable request to the authors

## ACKNOWLEDGEMENTS

The authors thank all participating healthcare professionals and Private Practitioners across the state and contributing Hospitals and institutions for their valuable contributions.We also acknowledge the support of research assistants and the state medical registrar in facilitating data collection.

